# A Nutrient Ratio–Based, Web-Enabled Food Quality Score Performs Favorably Compared with Leading Nutrient Profiling Systems for Weight and Blood Pressure

**DOI:** 10.1101/2024.12.23.24319328

**Authors:** Christopher J. Damman, Cara L. Frankenfeld

## Abstract

**Background:** Rising prevalence of chronic cardiometabolic conditions may be partly driven by shifts in dietary patterns. Nutrient profiling systems (NPSs) aim to guide healthier food choices through labeling and consumer-facing technologies but vary in accessibility and how well they distinguish food healthfulness.

**Objectives:** The primary objective was to compare a ratio-based, web-enabled NPS, Nutrient Consume Score (NCS), and its underlying nutrient ratios, with four leading NPSs—Nutri-Score (NS), Health Star Rating (HS), NOVA Classification (NC), and Food Compass 2.0 (FC)—by examining associations with obesity and blood pressure. Secondary objectives included assessing associations with cardiometabolic biomarkers and identifying food categories contributing most to each score.

**Methods:** NHANES 2015–2016 data for adults aged ≥20 years were analyzed. Dietary intake was assessed via 24-hour recalls, and NPS scores were calculated. Multivariable regression models adjusted for sociodemographic and health factors examined associations with obesity, blood pressure, and cardiometabolic biomarkers. Compositional analysis evaluated food categories driving the scores.

**Results:** The ratio-based NCS performed favorably compared with others. All NPSs significantly associated with healthier weight, while blood pressure associations were more variable. Including alcohol and adjusting for bioactives strengthened the ratio-based NCS’s association with weight outcomes. Across NPSs, food categories contributing to high and low scores were broadly consistent, generally reflecting minimally processed foods for high scores and more processed foods for low scores.

**Conclusions:** The Nutrient Consume Score (NCS) showed favorable associations with weight and blood pressure relative to other NPSs, highlighting its potential for improving diet-related risk factors for chronic conditions. As a web-enabled NPS, NCS may be well suited for consumer-facing technologies, such as smartphones, and could facilitate interventional trials to assess the causal impact of diet quality on cardiometabolic outcomes.

## Introduction

The rising incidence and prevalence of obesity (1), metabolic syndrome (2), and other noncommunicable disease (3) in the United States and globally (4) has been linked to shifts in the types of foods available to consumers including increased ultra-processed (5) and hyperpalatable foods (6) with resulting changes in dietary habits (7). Complex, confusing (8), and conflicting (9) nutrition guidance make it difficult for individuals to make informed, healthy choices on food purchases, preparation, and consumption (10).

Nutrient profiling systems (NPSs) aim to simplify nutrition complexity into easier-to-follow ratings that assess the health quality of foods and beverages. These systems can offer real-time guidance through package labeling (11), online websites (12), and smartphone technologies (13,14), helping individuals make healthier food choices. Additionally, they can support public health recommendations (15) and guide food companies in creating healthier products (16,17). While there are over 100 reported NPSs (15,18), a small subset has been rigorously evaluated for their association with health outcomes. The most studied NPSs with strongest links to improved dietary choices and better health are Nutri-Score (NS) (19), Health Star (HS) (20), NOVA Classification (NC) (21), and Food Compass (FC) (22,23).

Nutri-Score (24), popular in several European countries, and Health Star (25), widely used in Australia and New Zealand, rank foods based on nutrient profiles and food categories. The NOVA classification categorizes foods by their degree of processing, popularizing the term ultra-processed food (26). Food Compass evaluates foods across nine health domains, including categories like nutrient ratios, fiber, and phytochemicals (23,27). These systems are variably effective in assessing the relative quality of different ultra-processed foods (28) and not available on front of product packaging in many countries including the United States(23).

Despite the availability of multiple NPSs, a gap remains for a rigorously studied, web-enabled algorithm that is independent of country-specific labeling practices and that can scale, based on relative rather than absolute nutrient measures. The Nutrient Consume Score (NCS) (12) is a recently developed web-based algorithm that focuses on nutrient ratios – carbohydrate-to fiber (29), saturated fat-to-unsaturated fat (30), sodium-to-potassium (31), and calorie-to-weight (32) – as proxies for the degree of food processing and previously identified as strong predictors of food quality (33). It also incorporates alcohol and adjusts for food categories high in bioactive components that support a healthy gut microbiome, including polyphenols, bioactive fats, fermentable fibers, and fermentation products (34).

The primary aim of this study was to compare NCS, a nutrient ratio-based, web-enabled food quality score with other leading NPSs in relation to weight and blood pressure, key prognostic factors in cardiometabolic disease and mortality.

## Methods

### Data Sources and Analytic Population

The National Health and Nutrition Examination Survey (NHANES) is a repeated, cross-sectional analysis that has been conducted continuously since 1999 and data are released in two-year cycles, with exceptions for recent cycles due to COVID-19 disruptions in survey data collection(35). The survey includes interview and physical measurement components and is conducted in a nationally representative sample of approximately 5000 persons each year from persons located in 15 counties that are randomly selected each year.

For the primary analysis to compare NCS and other nutrient profiling scores, data from the continuous survey data releases 2015-2016 was used for this analysis because complete data was available for all evaluated NPS scores for this NHANES data cycle (36). In the 2015-2016 data cycle, 9971 individuals participated. The following exclusions were applied in this order to obtain the analytic subsample used (n excluded): age <20 years (n=4252), missing education (n=5), missing poverty-to-income ratio (n=0), missing day 1 dietary intake data (n=699), physiologically unsustainable low dietary intake of <500 kcal (n=42), physiologically unsustainable high dietary intake >5000 kcal (n=57), missing smoking information (n=20), missing waist circumference (n=203), missing body mass index (n=9), missing blood pressure (n=51), or missing physical activity (n=35). NHANES does not have missing data for age, gender, or race/ethnicity. The analytic sample for primary analyses used included 4598 adult individuals. A secondary analysis was undertaken to evaluate associations of the NCS score with other cardiometabolic biomarkers in NHANES cycles 2005-2018. Additional exclusions to the 4598 individuals in the analytic sample were missing LDL, HDL, or total cholesterol, plasma glucose, or blood triglycerides (n=2647). The analytic sample for secondary analyses of cardiometabolic biomarkers included 1951 adult individuals.

### Dietary Intake Assessment

Dietary intake is assessed for NHANES using two multiple pass 24-hour recalls. The first 24-hour recall is conducted in person during the visit to the Mobile Examination Center (MEC). The second 24-hour recall is conducted via phone 3 to 10 days after the MEC visit. All NHANES participants are eligible for the 24-hour recall interviews. The interviews are conducted using a midnight-to-midnight time frame for the 24-hour period prior to the interview. The USDA Food Survey Research Group conducts the dietary data collection methodology, maintenance of databases used to code and process data, and data review and processing. The dietary data is released in two files: individual foods and total nutrients. The individual foods file lists each food reported by the participant, along with details of the consumption such as a USDA FNDDS code (food code), eating occasion, and amount of food/beverage consumed in grams. The total nutrient intake file is a daily aggregate of nutrients from the reported foods consumed for that 24-hour recall.

### Nutrient Profiling Systems and Nutrient Ratios

This study evaluated five nutrient profiling systems, each rescaled to a 100-point scale for comparison. Nutri-Score (A–E = 100–20), Health Star Rating (0.5–5 = 10–100), NOVA classification (1–4 = 80–20), and Food Compass 2 (1–100 = 1–100) scores were converted from previously reported values (23,27). Alcoholic beverages were assigned null values in systems that did not evaluate them: NS, HS, NC, and FC. Nutrient Consume Score values were retrieved using the NCS algorithm available online at Gutbites.org (12). Additionally, nutrient ratios—carbohydrate-to-fiber, saturated fat-to-unsaturated fat, sodium-to-potassium, and calorie-to-weight—were mapped to a quantized scale of 1 to 3 in 0.125 increments before analysis.

Nutri-Score is a scoring algorithm that balances positively weighted nutrients (e.g., fruits, vegetables, fiber) against negatively weighted ones (e.g., saturated fat, sugars, sodium), resulting in a color-coded rating from A to E (24). Health Star Rating separates foods into three groups – dairy products, non-dairy beverages, and all other foods – and assigns points based on levels of protein, fats, saturated fats, energy, carbohydrates, sugar, and sodium, yielding in a 5-star scale with half-star increments (25). The NOVA Classification ranks foods by processing level, distinguishing between minimally or unprocessed foods, processed ingredients, processed foods, and ultra-processed foods, resulting in a 4-level scale (26). Food Compass assesses foods across nine domains – nutrient ratios, vitamins, minerals, food-based ingredients, additives, processing, specific lipids, fiber & protein, phytochemicals – producing a 100-point score (23,27).

Nutrient Consume Score (NCS) is an online nutrient profiling system based on nutrient ratios – Carbohydrate-to-Fiber, Saturated Fat-to-Unsaturated Fat, Sodium-to-Potassium, and Calorie-to-Weight – and other nutrients (protein, alcohol, vitamin D, iron, calcium) available on U.S. Nutrition Facts labels (12). Positive adjustments are made for food categories like fruits, vegetables, nuts, seeds, and whole grains to account for microbiome-supportive and bioactive factors not listed on food labels: polyphenols, prebiotic fibers, bioactive fats, and fermentation products. In addition, negative adjustments are made for alcohol, soft drinks, processed meats, and processed potatoes to reflect additives and food components linked to negative health outcomes. Three versions of NCS were evaluated in the analysis: NCS proper, NCS without alcohol included in calculations to compare to other algorithms that don’t include alcohol, and NCS without bioactive adjustments.

All scores and nutrient ratios were evaluated in three ways: weighted by calorie (kcal), weighted by weight (grams), and unweighted. Unweighted scores were calculated for each individual as the average score of each food consumed in the 24-hour period divided the total number of food items. Scores weighted by energy and weight were calculated as the sum of the individual food items’ scores multiplied by the kcal or grams provided by the food item and then divided by the total kcal or grams for the 24-hour period. Scores were analyzed as continuous variables on a 10-unit scale.

### Food Categories and Contributions to Scores

To evaluate which foods were contributing to most of the variation in the scores, a compositional approach was applied. Individual foods reported were assigned to food group classifications based on the What We Eat in America (WWEIA) food categories (37). Total intake of food categories was calculated for each person based on percentage energy (kcals) provided by all the foods in that category for the 24-hour dietary intake recall period. Center-log transformed ratios of percentage energy contributed by food categories to overall energy were calculated (38) using the R *compositions* package (39) to include in regression models.

### Outcomes

Anthropometry and blood pressure were evaluated as continuous outcomes and as binary outcomes based on established cut-offs. Anthropometric characteristics are measured during the in-person visit by trained technicians. Blood pressure was measured one to four times by trained technicians. Details about measurement and analysis are available from NHANES established procedures. The average of the available measurements for each person was used for analysis.

Obesity was defined as BMI 30 kg/m^2^ (40) and abdominal obesity was defined as waist circumference >88 cm for women and >102 cm for men (41). High blood pressure was defined as the presence of systolic blood pressure 130 mm/Hg or diastolic blood pressure 85 mm/Hg (41,42).

Other cardiometabolic risk factors (comparison with NCS only) evaluated included low-density serum lipoprotein (LDL), high-density lipoprotein (HDL), total cholesterol, and triglycerides, and plasma glucose. These risk factors were evaluated as continuous measures and in relation to established cut-offs for high or low values (41,42). Additionally, metabolic syndrome was assessed and was classified as the presence of three or more of abdominal obesity, high blood pressure, high LDL, low HDL, or high serum glucose (41).

### Covariates

Personal characteristics were included to describe the analytic group and as adjustment variables in multivariable analyses. Characteristics included were: age (in categories for description and in years for multivariable analyses), gender (female and male), race or ethnicity (non-Hispanic White, non-Hispanic Black, non-Hispanic other, and Hispanic). Education (< high school graduate; high school graduate or equivalent; some college or associate degree; and, four-year college graduate or more), poverty-to-income ratio (PIR, <1, 1 to <2, 2, to <3, and 3+), smoking status (less than 100 cigarettes in lifetime; former smoker; current <20 cigarettes per day; and, current 20+ cigarettes per day), and physical activity (<15, 15 to <75, 75 to <165, 165+ metabolic equivalent score (METS) per week).

### Statistical Analysis

For visualization of overall differences in relation to increasing NCS score, descriptive statistics for covariates were calculated by tertiles of the energy-weighted NCS score. Means and standard errors were calculated for continuous variables and frequencies and percentages were calculated for categories variables. To evaluate associations of NPSs and ratio scores with anthropometry, blood pressure, and cardiometabolic biomarkers, multivariable linear and logistic regression analyses were performed for continuous and binary outcomes, respectively, and adjusted for personal characteristics (age, gender, race/ethnicity, education, PIR, smoking, and exercise. For food category contributions to scores, linear regression analyses were used to evaluate the contribution of center-log ratio transformed food categories of the energy-weighted scores and subscores. Except for the compositional analysis of food category contributions to NPSs and ratios scores, analyses were conducted using population weights supplied by NHANES and incorporating the complex survey design. A subpopulation command to maintain appropriate population weights for the study population with complete data for this analysis.

Statistical significance was considered p<0.05 for analyses with cardiometabolic and mortality outcomes. Statistical analysis was conducted in Stata (StataCorp, TX, USA, version 18).

## Results

### Study Population Characteristics

For visualization, study population characteristics by tertiles of energy-weighted NCS scores are presented (Table 1). Categories more represented in the highest score tertiles included age greater than 65, female gender, Hispanic, non-Hispanic other, four-year college graduates, poverty-to-income ratio of three or higher, and never smokers. Categories more represented in the lowest score tertiles included younger age, non-Hispanic White, non-Hispanic Black, lower education level, current smoker, and highest physical activity level (165+ METS).

**Table 1.** Study population characteristics by tertiles of energy-weighted NCS score in NHANES adults, 2015-2016.

| Personal characteristics | Tertile 1 (<37.08),<br>n=1533 | Tertile 2 (37.08 to<br>48.31), n=1533 | Tertile 3 (>48.31),<br>n=1532 |
| --- | --- | --- | --- |
| Age (years) |  |  |  |
| 20 to 34 | 458 (33.3%) | 370 (25.4%) | 360 (25.4%) |
| 35 to 49 | 415 (26.8%) | 392 (24.8%) | 339 (22.6%) |
| 50 to 64 | 380 (25.2%) | 434 (28.6%) | 403 (27.4%) |
| 65+ | 280 (14.8%) | 337 (21.2%) | 430 (24.7%) |
| Gender |  |  |  |
| Male | 764 (53.3%) | 752 (47.1%) | 693 (43.0%) |
| Female | 769 (46.7%) | 781 (52.9%) | 839 (57.0%) |
| Race and non-Hispanic ethnicity |  |  |  |
| Non-Hispanic White | 589 (67.7%) | 559 (68.0%) | 420 (58.2%) |
| Non-Hispanic Black | 415 (13.8%) | 316 (10.4%) | 220 (7.6%) |
| Non-Hispanic Other | 116 (4.9%) | 199 (8.2%) | 333 (16.2%) |
| Hispanic | 413 (13.6%) | 459 (13.4%) | 559 (18.0%) |
| Education |  |  |  |
| <HS graduate or equivalent | 293 (12.4%) | 324 (12.8%) | 394 (13.5%) |
| HS graduate or equivalent | 412 (26.9%) | 347 (21.6%) | 264 (13.8%) |
| Some college or associates degree | 527 (36.1%) | 463 (33.4%) | 389 (30.6%) |
| Four-year college graduate or more | 301 (24.6%) | 399 (32.2%) | 485 (42.2%) |
| Poverty-to-income ratio |  |  |  |
| <1 | 307 (14.7%) | 265 (10.2%) | 312 (13.5%) |
| 1 to <2 | 399 (19.4%) | 387 (20.5%) | 338 (15.0%) |
| 2 to <3 | 252 (18.0%) | 262 (16.9%) | 219 (12.8%) |
| ≥3 | 460 (41.6%) | 488 (45.8%) | 500 (49.8%) |
| Missing | 115 (6.3%) | 131 (6.6%) | 163 (8.9%) |
| Smoking |  |  |  |
| Never | 807 (51.6%) | 869 (55.4%) | 1,009 (64.1%) |
| Former | 338 (23.7%) | 360 (26.7%) | 365 (25.4%) |
| Current, less than 20 cig/day | 282 (16.0%) | 242 (14.2%) | 139 (8.6%) |
| Current, 20+ cig/day | 106 (8.6%) | 62 (3.7%) | 19 (2.0%) |
| Physical activity (METS/week) |  |  |  |
| <15 | 666 (38.2%) | 680 (40.4%) | 688 (36.1%) |
| 15 to <75 | 431 (31.3%) | 453 (32.7%) | 510 (41.2%) |
| 75 to <165 | 219 (15.8%) | 195 (14.4%) | 197 (13.6%) |
| 165+ | 217 (14.7%) | 205 (12.5%) | 137 (9.0%) |

### Weight and Blood Pressure Associations with Scores and Ratios

All energy-weighted NPS scores (NCS, FC, HS, NC and NS) were significantly associated with continuous measures of anthropometry and blood pressure (Figure 1). Similar associations were observed with grams-weighted and unweighted scores (Supplementary Table 1). NCS and FC showed the largest beta-coefficients for BMI and waist circumference. NCS without alcohol and NS showed the largest beta-coefficients for systolic and diastolic blood pressure. Removing bioactives from NCS scoring reduced beta-coefficients for association with anthropometry and blood pressure. Removing alcohol from NCS scoring reduced beta-coefficients for association with anthropometric measures, but had the opposite effect on blood pressure.

**Figure 1.**
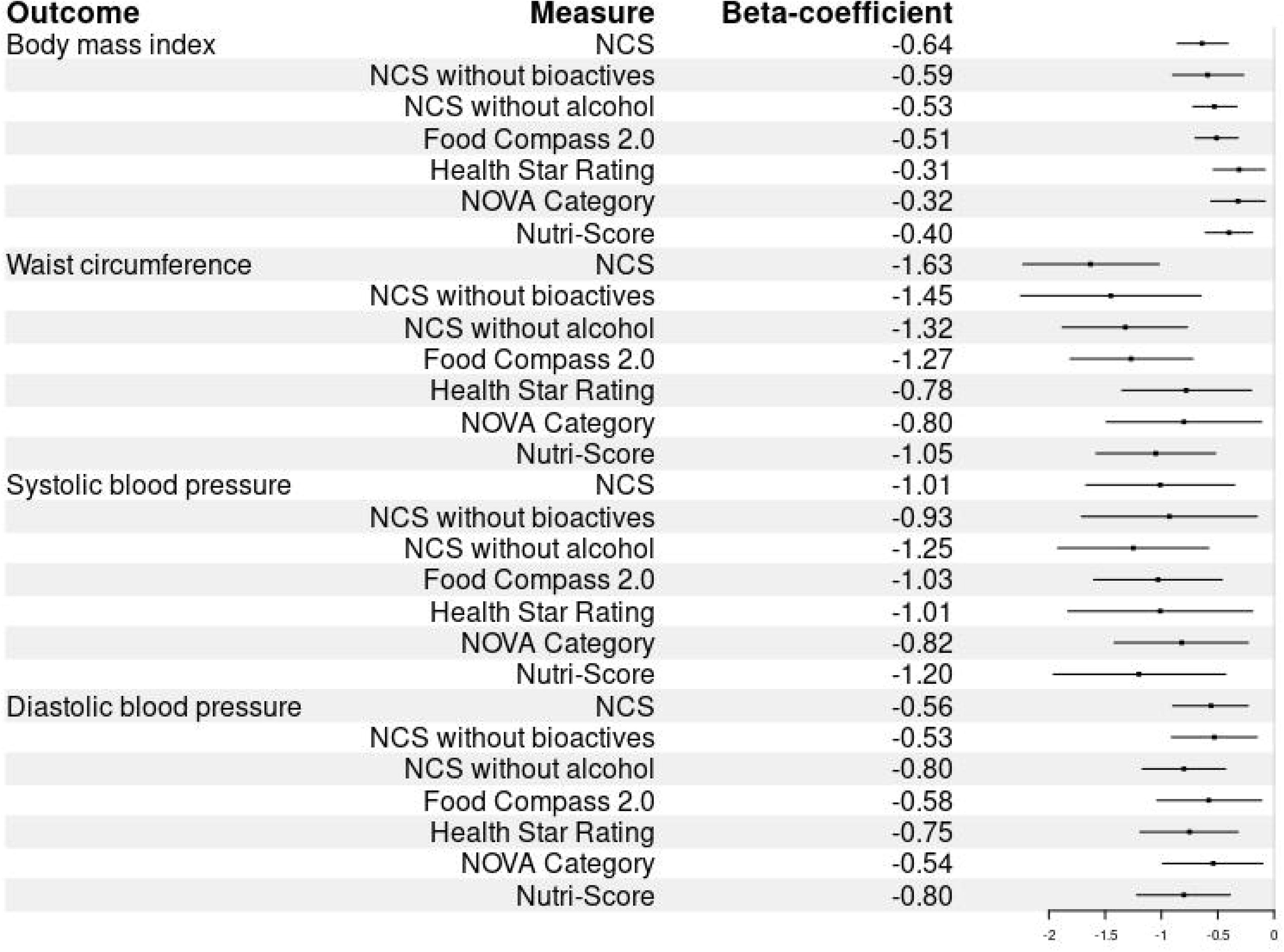
Associations of nutrient profile scores (in 10-unit values) with body mass index, waist circumference, and blood pressure. Beta-coefficients from multivariable adjusted models are displayed. Models are adjusted for age, gender, race or Hispanic ethnicity, education, smoking, poverty-income ratio, and physical activity.

Higher scores for all energy-weighted NPS scores (NCS, FC, HS, NC, and NS) were associated with lower odds of obesity, abdominal obesity, and high blood pressure (Figure 2), with similar associations observed with grams-weighted and unweighted scores (Supplementary Table 1). NCS and FC showed the lowest odds for obesity, while NCS and NS showed the lowest odds for abdominal obesity. NCS without alcohol and NS showed the lowest odds for high blood pressure. Removing bioactives or from NCS scoring reduced odds of obesity, abdominal obesity, and high blood pressure. Removing alcohol from NCS scoring reduced odds of obesity, but had the opposite effect on odds of high blood pressure.

**Figure 2.**
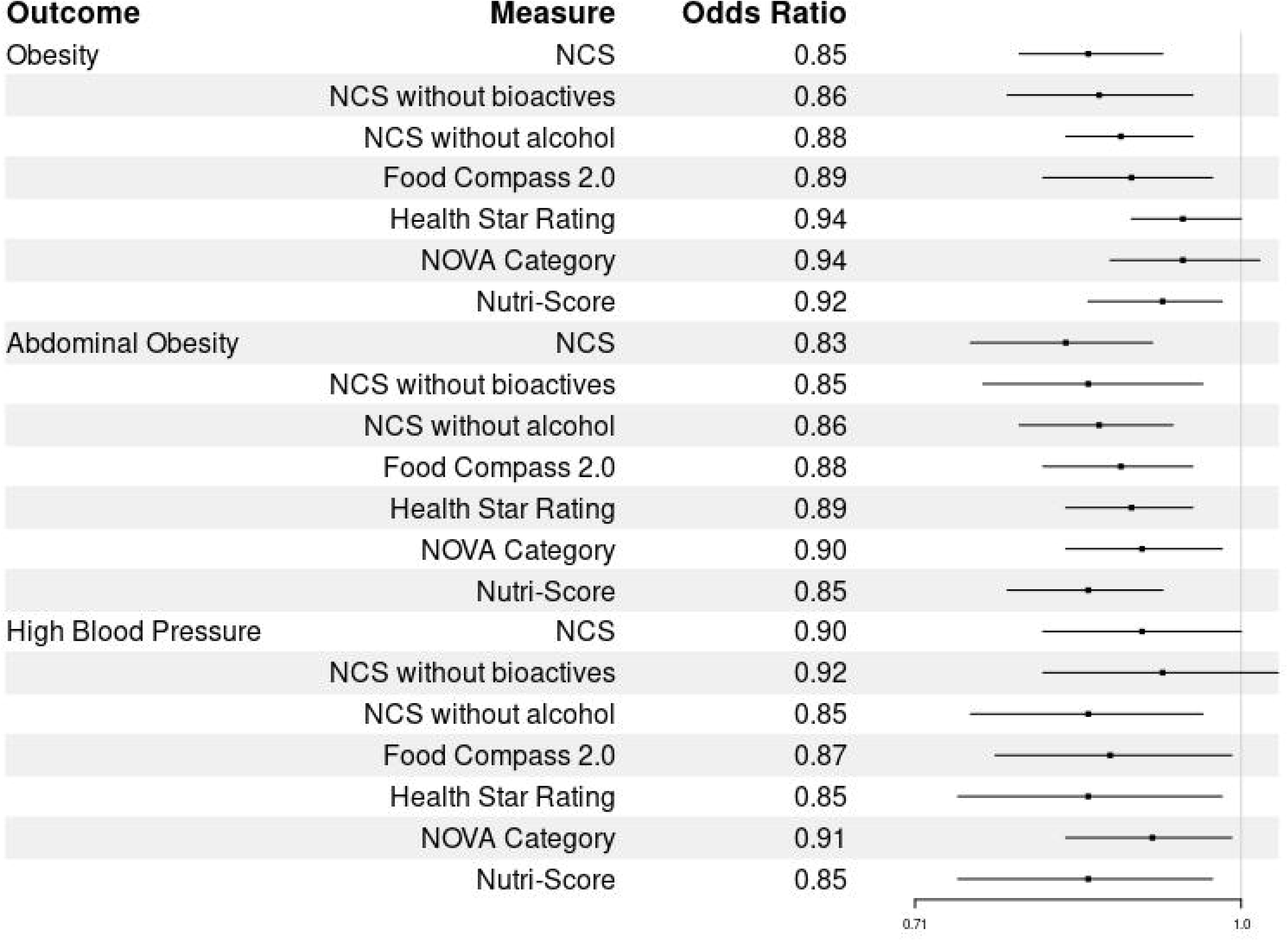
Associations of nutrient profile scores (in 10-unit values) with obesity, abdominal obesity, and high blood pressure. Odds ratios from multivariable adjusted models are displayed. Models are adjusted for age, gender, race or Hispanic ethnicity, education, smoking, poverty-income ratio, and physical activity.

Nutrient ratio scores (energy, salt, fat, fiber; Figure 3 and Figure 4) accounted for much of the signal observed across the full scoring systems, with outcome-specific effects. Ratios were scaled such that higher values reflected less favorable nutrient profiles. Energy, salt and fat ratios were favorably associated with anthropometric measures; while associations with blood pressure varied: salt and fiber ratios were favorable, while energy ratios were unfavorable.

**Figure 3.**
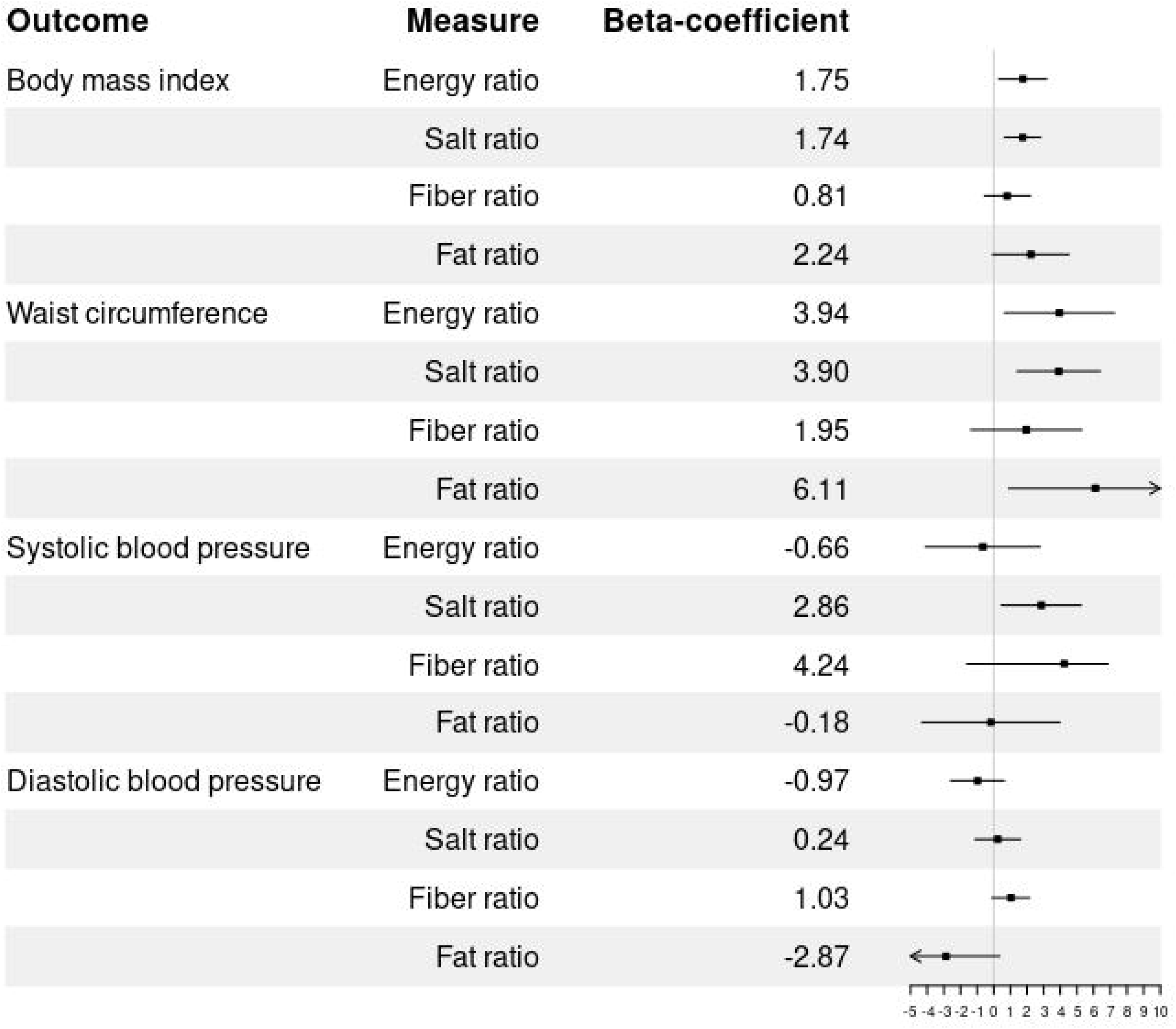
Associations of nutrient ratio scores (in 1-unit values) with body mass index, waist circumference, and blood pressure. Beta-coefficients from multivariable adjusted models are displayed. Models are adjusted for age, gender, race or Hispanic ethnicity, education, smoking, poverty-income ratio, and physical activity.

**Figure 4.**
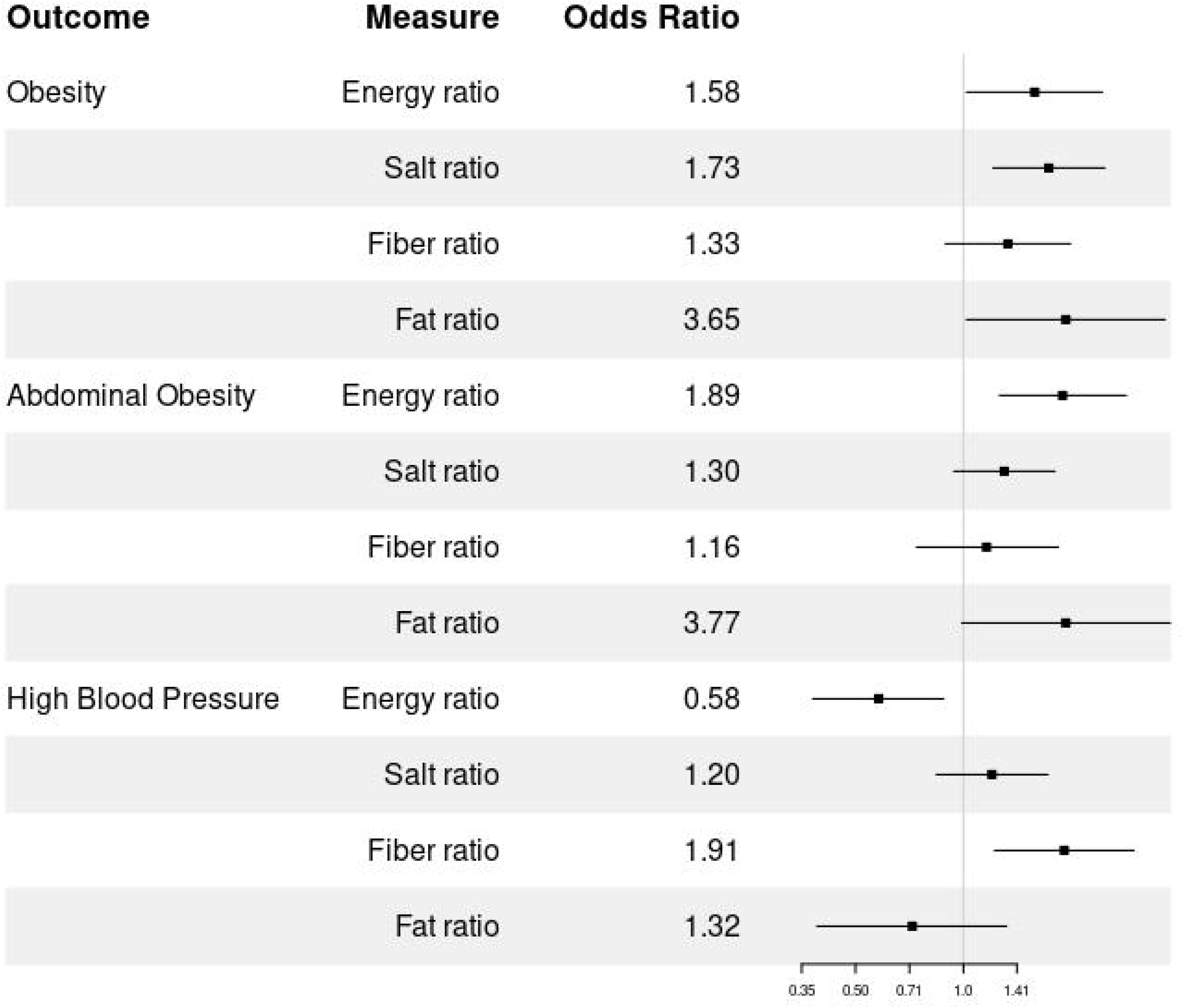
Associations of nutrient ratio scores (in 1-unit values) with obesity, abdominal obesity, and high blood pressure. Odds ratios from multivariable adjusted models are displayed. Models are adjusted for age, gender, race or Hispanic ethnicity, education, smoking, poverty-income ratio, and physical activity.

### Other Cardiometabolic Risk Factor Associations with Scores and Ratios

Among cardiometabolic risk factors (Supplemental Figures 2A, 2B, 3A, and 3B), there was an association between NCS, FC and higher total cholesterol. Fiber ratio was favorably associated with higher HDL and salt ratio was favorably associated with lower glucose.

### Food Category Contributors to Scores

The top food categories contributing to NPS variation were largely consistent across scores, (Figure 5, Supplemental Figure 4). Among negatively contributing categories, NCS provided relatively higher weight than most other NPS’s to “pizza”, “soft drinks”, “frankfurters” and “egg/breakfast sandwiches”. Among positively contributing categories, NCS provided relatively higher weight than most other NPS’s to “Rice”, “Beans, peas, and legumes”, “nuts and seeds”. A summary of NCS food scores within different categories can be found in Supplemental Figure 1.

**Figure 5.**
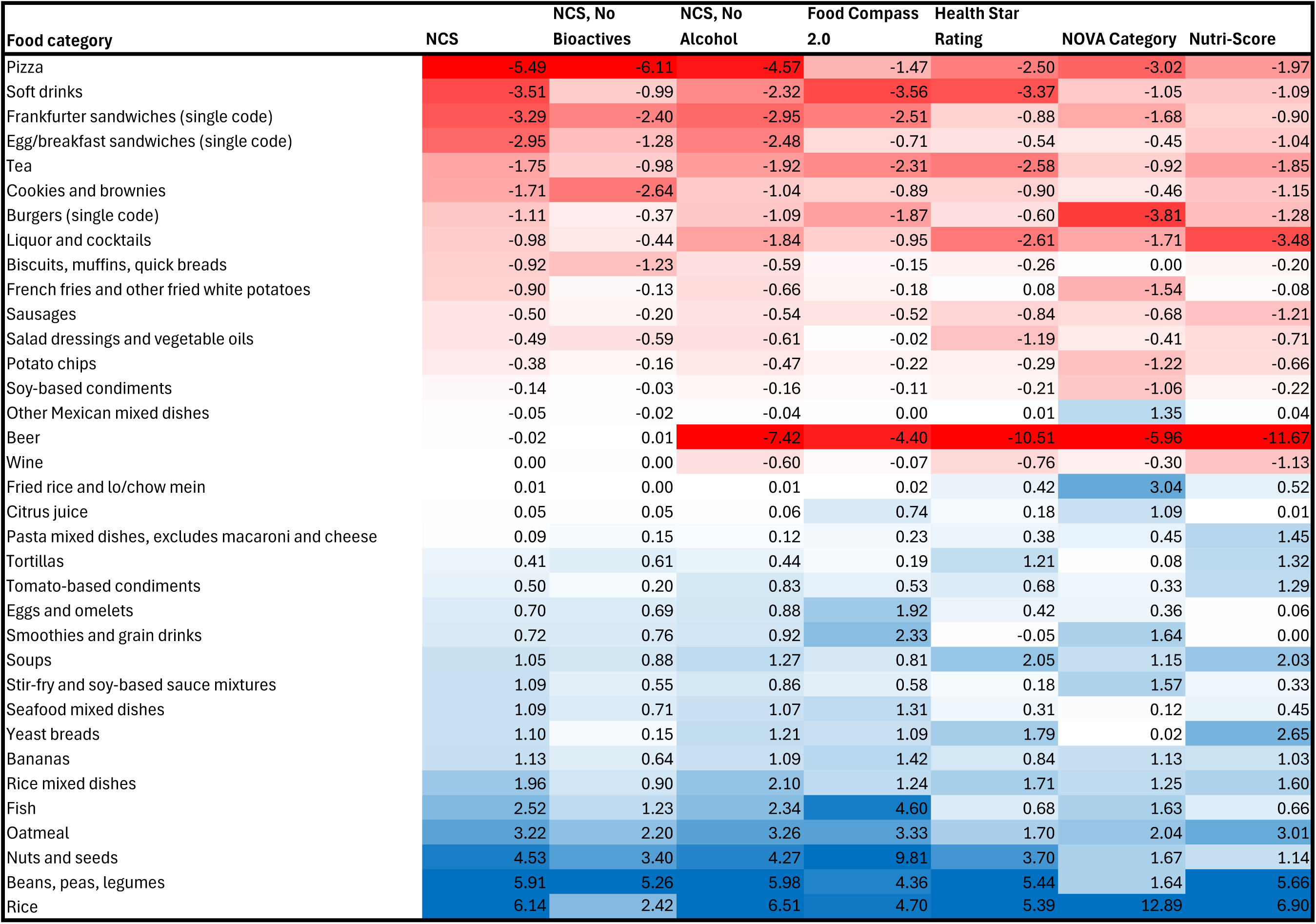
Heatmap of food category contributions to kcal-weighted scores. Values represented are the percentage of variation explained (R-squared*100*indicator for direction of association). Food categories that had no consumption in the study population and foods not included in a WWEIA category are not included, and only foods for which at least it explains at least 1% of variation of a score are included. Darker shading represents more variation explained, and red indicates inverse associations and blue indicates positive associations.

## Discussion

Nutrient profiling systems assess the quality of food and beverages, and aim to help consumers, industry, and government select, create, and promote healthier options to improve public and individual health (18). This study shows that leading NPSs are associated with weight and blood pressure, key prognostic factors in cardiometabolic disease, neurological disorders, cancer, and mortality. A ratio-based web-available score performed favorably relative to leading NPSs and also associated with a higher cholesterol likely driven by high favorable HDL. Notably, the food categories contributing greatest to scores were largely consistent across systems. Those foods contributing most to positive scores included minimally processed foods and those that contributed most to negative scores included ultra-processed foods.

A key finding is that much of the power of NPSs may be captured in the following nutrient ratios: carbohydrate-to fiber (29), saturated fat-to-unsaturated fat (30), sodium-to-potassium (31), and calorie-to-weight (32), which all associated with weight and variably associated with blood pressure. Nutrient ratios reflect nutritional balance in whole foods and the degree to which certain nutrients are concentrated or depleted in processed foods. Ratios are explicitly incorporated in systems like NCS and FC and implicitly applied in systems like NS and HS, which assign positive and negative weights to under- and overrepresented nutrients. Nutrient ratios may complement the NOVA classification, which has been critiqued for grouping all ultra-processed foods together, by offering an empirical approach to assessing the relative healthfulness of different ultra-processed foods (43). The unfavorable association of calorie-to-weight with blood pressure could be related to beverages (including juices and sodas) having lower nutrient density and future work may benefit from treating nutrient density of beverages and foods differently.

Another key finding is that including alcohol in a ratio-based score may enhance its health-discriminatory power for weight, but not blood pressure, perhaps also owing to complexities around beverages. In NPSs that excluded alcohol from their calculations, alcoholic beverages contributed to the scores, likely due to co-consumption of alcohol with other foods. This result also highlights an important point about the role of alcohol in overall dietary intake for individuals that choose to consume alcohol. Alcohol contributes at least 16% of the average US adult’s daily caloric intake (44), a figure that is likely higher since the impact of COVID-19 on consumption (45). However, many are unaware of alcohol’s impact (46). Given the prevalence of alcohol consumption and its association with obesity (47) and other adverse health outcomes(48), there is a need for consumer education and NPSs that account for alcohol’s caloric contribution to encourage healthier dietary choices.

A third key takeaway is that removing adjustments for food groups serving as proxies for bioactive factors diminished the ability of NCS to discriminate associations with weight and blood pressure measures. These food group adjustments included both those linked to positively associated microbiome-active and bioactive factors (e.g., phytochemicals, fermentable fibers, and bioactive fats) and those linked to negatively associated bioactives (e.g., alcohol, fructose, nitrites, acrylamide, advanced glycosylation end products, and trans fats). A better understanding and measurement of these positive and negative health-associated bioactive factors, along with their explicit inclusion in food labels and NPS algorithms, are key focuses of current nutrition research (34).

NPSs have been shown to positively influence consumer buying habits (49) and prospective cohort studies have shown they can positively impact health measures like obesity and mortality (19,50). They can play a key role in public health messaging through front-of-package labels (11). In countries where front of package labels don’t yet exist, web-enabled portals (12), and smartphone technologies that scan product UPCs to determine food quality could be useful (13,14). A ratio-based score in particular could intuitively encourage individuals to incorporate more fiber-, potassium-, unsaturated fat-rich foods, potentially countering the health impact of foods high in simple carbohydrates, sodium, and saturated fat. A ratio-based NPS could also guide the food industry in developing healthier products by providing empirical guide rails to rebalance simple carbohydrates with fiber (51,52) and sodium with potassium (53).

Nutrient profiling systems (NPSs) hold promise for improving health outcomes, but they have limitations. Some systems generate outlier results for certain foods that conflict with epidemiological data (54). Most exclude alcohol, despite its strong association with adverse health outcomes (19,27,55), while others are critiqued for being overly simple or too complex (22). Additionally, most NPSs rely on nutritional databases that lack quantification of the bioactive components of foods, such as microbiome-active components (i.e. polyphenols, fermentable fibers, and fermentation products), and are missing data on food additives that may harm the microbiome and individual (56). Most NPSs fail to account for a food’s matrix or ultra-structure, which may be important for understanding the health benefits of unprocessed foods (57). NPS algorithms don’t currently provide personalized advice which may be important to account for individual variations in nutrient requirements, including those influenced by the gut microbiome (58,59). Tailoring algorithms in low- and middle-income country cohorts, where baseline macro- and micronutrient intakes differ significantly, may also be necessary.

This study design also has limitations including selection (60), recall (61), and reporting (62) biases. While NHANES attempts to survey respondents to obtain representativeness to the US population, there is still self-selection of participants who are invited to participate in NHANES, and individuals may misreport information in surveys and interviews due to misremembering or aligning responses with social desirability. Additionally, the single assessment of dietary intake may not be reflective of longer term or usual intake. However, the large sample size of NHANES does mitigate some of these biases. Temporality of associations cannot be determined due to the cross-sectional design of NHANES, and weight measures in particular can be subject to reverse causation bias (63). Results regarding weight should be interpreted in this context, and it is possible that individuals of different body sizes eat differently rather than the scores predict weight. Cohort studies demonstrating prospective association between Nutri-score and abdominal obesity suggest that reverse causation may not entirely explain the associations observed (19).

In conclusion, this study highlights the potential of nutrient profiling systems (NPSs) including a nutrient ratio-based web-enabled score to support personal and public health efforts in curbing weight gain and improving blood pressure. Ratio-based systems, in particular, may guide individuals, food companies, and governments in rebalancing nutrients in our diet. As a ratio-based system, NCS may be particularly useful at measuring the spectrum of health of different processed and ultra-processed foods (Supplementary Figure 1). A ratio-based system also has advantages in being independent of absolute values of nutrients so it operates independent of scale and allows assessing food combinations. While NPSs offer valuable insights, most don’t account for alcohol and are primarily supported by correlative research. NCS also offers an advantage over other algorithms given its web accessibility. Future studies leveraging smartphone access to the web-enabled algorithm could assess NCS’s capacity for personalization, generalizability across diverse populations, and causal effects on metabolic health.

## Data Availability

All data produced in the present work are contained in the manuscript

## Acknowledgements

Grateful to Luke Walker for his vital assistance with coding the scoring algorithms and to Ben Roberts for his invaluable feedback and constructive criticism on the manuscript. CJD designed and conducted the research, drafted the paper, and was responsible for final content. CLF performed the statistical analysis, wrote the methods section, and revised the manuscript. All authors have read and approved the final version.

## Data Availability

NHANES data used in this work is freely available from the National Center for Health Statistics. Other data described in the manuscript and a code book will be made available upon request pending approval.

## Declaration of Generative AI and AI-assisted technologies in the writing process

During the preparation of this manuscript, ChatGPT was used on a limited basis to improve clarity and wording of the text. The authors subsequently reviewed and revised all content. No generative AI tools were used for data analysis or interpretation.

## Figure Legends

**Supplemental Figure 1.** Distribution of NCS scores for representative foods, organized by food category.

**Supplemental Figure 2.** Associations of nutrient profile scores (in 10-unit values) with high LDL cholesterol, low HDL cholesterol, high total cholesterol, high triglycerides, and high plasma glucose. Beta-coefficients for continuous outcomes (panel A) and odds ratios for dichotomous outcomes (panel B) from multivariable adjusted models are displayed. Models adjusted for age, gender, race or Hispanic ethnicity, education, smoking, poverty-income ratio, and physical activity.

**Supplemental Figure 3.** Associations of nutrient ratios (in 1-unit values) with LDL cholesterol, HDL cholesterol, total cholesterol, triglycerides, and plasma glucose. Beta-coefficients for continuous outcomes (panel A) and odds ratios for dichotomous outcomes (panel B) from multivariable adjusted models are displayed. Models are adjusted for age, gender, race or Hispanic ethnicity, education, smoking, poverty-income ratio, and physical activity.

**Supplemental Figure 4.** Heatmap of food category contributions to kcal-weighted scores. Values represented are the percentage of variation explained (R-squared*100*indicator for direction of association). All WWEIA food categories displayed. Darker shading represents more variation explained, and red indicates inverse associations and blue indicates positive associations.

**Supplemental Table 1.** Associations of nutrient profile scores (in 10-unit values) with continuous measures of anthropometry and blood pressure, and with dichotomous measures for obesity, abdominal obesity, and high blood pressure, using unweighted, weighted by grams, and weighted by kcal approaches to scores.

|  | NCS | Food Compass 2.0 | Health Star Rating | Nova Category | Nutri-Score |
| --- | --- | --- | --- | --- | --- |
| Multivariable regression results | beta/OR (CI), p-value | beta/OR (CI), p-value | beta/OR (CI), p-value | beta/OR (CI), p-value | beta/OR (CI), p-value |
| **Unweighted** |  |  |  |  |  |
| Body Mass Index (kg/m<sup>2</sup>) | -0.86 (-1.18,-0.55), <0.0001 | -0.73 (-1.02,-0.43), 0.0001 | -0.72 (-1.05,-0.39), 0.0003 | -0.97 (-1.39,-0.55), 0.0002 | -0.78 (-1.13,-0.44), 0.0002 |
| Waist Circumference (cm) | -2.08 (-2.91,-1.26), 0.0001 | -1.73 (-2.48,-0.98), 0.0002 | -1.71 (-2.56,-0.87), 0.0006 | -2.32 (-3.41,-1.23), 0.0004 | -1.91 (-2.81,-1.01), 0.0004 |
| Systolic blood pressure (mmHg) | -0.65 (-1.28,-0.03), 0.0426 | -0.69 (-1.14,-0.24), 0.0055 | -0.73 (-1.53, 0.07), 0.0709 | -0.66 (-1.40, 0.07), 0.0729 | -0.84 (-1.66,-0.03), 0.0439 |
| Diastolic blood pressure (mmHg) | -0.30 (-0.65, 0.05), 0.0899 | -0.27 (-0.69, 0.15), 0.1915 | -0.52 (-0.86,-0.19), 0.0046 | -0.37 (-0.88, 0.13), 0.1382 | -0.49 (-0.89,-0.08), 0.0211 |
| Obese | 0.78 (0.71, 0.85), <0.0001 | 0.81 (0.73, 0.89), 0.0004 | 0.83 (0.75, 0.91), 0.0009 | 0.77 (0.68, 0.87), 0.0004 | 0.82 (0.74, 0.91), 0.0010 |
| Abdominal obesity | 0.81 (0.72, 0.92), 0.0034 | 0.83 (0.74, 0.93), 0.0032 | 0.82 (0.73, 0.92), 0.0023 | 0.76 (0.66, 0.87), 0.0009 | 0.81 (0.72, 0.91), 0.0015 |
| High blood pressure | 0.94 (0.86, 1.03), 0.1940 | 0.92 (0.84, 1.02), 0.1106 | 0.89 (0.80, 0.99), 0.0347 | 0.95 (0.85, 1.07), 0.4175 | 0.90 (0.80, 1.02), 0.0813 |
| **Weighted by grams** |  |  |  |  |  |
| Body Mass Index (kg/m<sup>2</sup>) | -1.00 (-1.29,-0.72), <0.0001 | -1.15 (-1.49,-0.80), <0.0001 | -0.86 (-1.14,-0.59), <0.0001 | -0.89 (-1.22,-0.57), <0.0001 | -0.70 (-0.93,-0.47), <0.0001 |
| Waist Circumference (cm) | -2.42 (-3.18,-1.66), <0.0001 | -2.69 (-3.58,-1.79), <0.0001 | -2.01 (-2.76,-1.26), <0.0001 | -2.10 (-2.94,-1.27), 0.0001 | -1.67 (-2.27,-1.07), <0.0001 |
| Systolic blood pressure (mmHg) | -0.04 (-0.63, 0.56), 0.9005 | -0.52 (-1.18, 0.15), 0.1190 | -0.39 (-1.01, 0.23), 0.1963 | -0.31 (-0.98, 0.36), 0.3419 | -0.33 (-0.90, 0.23), 0.2244 |
| Diastolic blood pressure (mmHg) | -0.05 (-0.35, 0.24), 0.7076 | -0.50 (-0.87,-0.12), 0.0124 | -0.41 (-0.69,-0.14), 0.0059 | -0.35 (-0.65,-0.06), 0.0233 | -0.26 (-0.50,-0.03), 0.0269 |
| Obese | 0.75 (0.69, 0.83), <0.0001 | 0.73 (0.64, 0.83), 0.0001 | 0.80 (0.73, 0.87), 0.0001 | 0.79 (0.70, 0.88), 0.0004 | 0.83 (0.78, 0.90), 0.0001 |
| Abdominal obesity | 0.76 (0.70, 0.84), <0.0001 | 0.74 (0.66, 0.83), <0.0001 | 0.79 (0.71, 0.88), 0.0003 | 0.79 (0.70, 0.89), 0.0009 | 0.82 (0.75, 0.91), 0.0008 |
| High blood pressure | 1.06 (0.98, 1.15), 0.1485 | 0.95 (0.85, 1.07), 0.4076 | 0.98 (0.90, 1.06), 0.5896 | 1.00 (0.92, 1.07), 0.8928 | 0.99 (0.92, 1.06), 0.7751 |
| **Weighted by kcal** |  |  |  |  |  |
| Body Mass Index (kg/m<sup>2</sup>) | -0.64 (-0.86,-0.41), <0.0001 | -0.51 (-0.70,-0.32), <0.0001 | -0.31 (-0.54,-0.08), 0.0124 | -0.32 (-0.56,-0.08), 0.0128 | -0.40 (-0.61,-0.19), 0.0012 |
| Waist Circumference (cm) | -1.63 (-2.23,-1.02), <0.0001 | -1.27 (-1.81,-0.72), 0.0002 | -0.78 (-1.35,-0.20), 0.0114 | -0.80 (-1.49,-0.11), 0.0253 | -1.05 (-1.58,-0.52), 0.0007 |
| Systolic blood pressure (mmHg) | -1.01 (-1.67,-0.35), 0.0051 | -1.03 (-1.60,-0.46), 0.0015 | -1.01 (-1.83,-0.19), 0.0187 | -0.82 (-1.42,-0.23), 0.0100 | -1.20 (-1.96,-0.43), 0.0045 |
| Diastolic blood pressure (mmHg) | -0.56 (-0.90,-0.23), 0.0026 | -0.58 (-1.04,-0.11), 0.0193 | -0.75 (-1.19,-0.32), 0.0022 | -0.54 (-0.99,-0.10), 0.0205 | -0.80 (-1.22,-0.39), 0.0009 |
| Obese | 0.85 (0.79, 0.92), 0.0004 | 0.89 (0.81, 0.97), 0.0138 | 0.94 (0.89, 1.00), 0.0359 | 0.94 (0.87, 1.02), 0.1117 | 0.92 (0.85, 0.98), 0.0207 |
| Abdominal obesity | 0.83 (0.75, 0.91), 0.0007 | 0.88 (0.81, 0.95), 0.0021 | 0.89 (0.83, 0.95), 0.0024 | 0.90 (0.83, 0.98), 0.0210 | 0.85 (0.78, 0.92), 0.0007 |
| High blood pressure | 0.90 (0.81, 1.00), 0.0586 | 0.87 (0.77, 0.99), 0.0399 | 0.85 (0.74, 0.98), 0.0297 | 0.91 (0.83, 0.99), 0.0333 | 0.85 (0.74, 0.97), 0.0235 |

