## Supplemental Figure 1 for "A Nutrient Ratio–Based, Web-Enabled Food Quality Score Performs Favorably Compared with Leading Nutrient Profiling Systems for Weight and Blood Pressure"

### **NUTRIENT CONSUME SCORE** at a Glance

| Color | Vegetables | Fruits | Nuts/Seeds/Beans | Savory Snacks | Sweet Treats | Cereals | Other Grains | Dairy/Milk | Meats/Fish/Egg | Meals | Cond/ Sauces | Beverages |
|---|---|---|---|---|---|---|---|---|---|---|---|---|
| >=90 | Kale (100)<br>Broccoli (100)<br>Carrots (100)<br>Tomatoes (100) | Raspberries (100)<br>Strawberries (100)<br>Orange (98)<br>Blueberries (94) | Lima Beans (98)<br>Edamame (95)<br>Lentils (94)<br>Black Beans (93) | | | | | | Fresh Tuna (96)<br>Fresh Salmon (90) | Fish Kabob (94)<br>Bouillabaisse (92) | Natto (92)<br>Marinara (91) | Water (100)<br>Celery Juice (93)<br>Black Coffee (90)<br>Green Tea (90) |
| >=80 | Cooked Veggies (80's)<br>Sweet Potato (89)<br>Kimchi (87)<br>Corn (86) | Cherries (88)<br>Plum (87)<br>Apple (86)<br>Cantaloupe (81) | Tofu (88)<br>Chia Seeds (84)<br>Flax Seeds (83)<br>Almonds (80) | | | Plain Oatmeal (89) | Injera (83)<br>Bulgar Wheat (82) | Greek Yogurt (86)<br>Ricotta Cheese (83)<br>Low Fat Yogurt (81)<br>Whole Fat Yogurt (80) | Canned Salmon (89)<br>Canned Sardines (89)<br>Baked Tilapia (84)<br>Grilled Salmon (83) | Beans & Rice (89)<br>Bean Soup (88)<br>Fish Curry (87)<br>Vegetable Curry (84) | Marinara w/ Meat (89)<br>Picco de Gallo (88)<br>Bean Dip (80)<br>Guacamole (80) | Beet Juice (88)<br>Veg Juice (87)<br>Protein Slimfast (86)<br>Muscle Milk (85) |
| >=70 | Winter Squash (77)<br>Rhubarb (75)<br>Sweet Pot. Fries (74)<br>Dill Pickles (72) | Dried Peach (79)<br>Mango (79)<br>Melon (75)<br>Watermelon (72) | Pecans (78)<br>Peanuts (76)<br>Cashews (70)<br>Peanut Butter (70) | | | Instant Oatmeal (76)<br>Cream of Wheat (73)<br>Shredded Wheat (70) | Multigrain Bread (78)<br>Quinoa (77)<br>Roti Bread (74)<br>Corn tortilla (71) | Cottage Cheese (79)<br>Soy Milk (79)<br>Low Fat Milk (78)<br>Whole Milk (77) | Chicken Breast (79)<br>Whole Egg (78)<br>Lean Beef (74)<br>Ground Chicken (70) | Tuna Sushi (76)<br>Chicken Curry (74)<br>Beef Stew (74)<br>Taco Salad w/ Meat (71) | Salsa (79)<br>Taco Sauce (76)<br>Hot Sauce (74)<br>Vodka Sauce (74) | Veg. Smoothie (79)<br>Nonfat Latte (77)<br>Non Alc. Wine (76)<br>Fruit Smoothie (75) |
| >=60 | Potato (69)<br>Creamed Spinach (67)<br>Tzatziki Dip (65)<br>Creamed Corn (60) | Canned Peach (69)<br>Pineapple (63)<br>Grapes (62)<br>Banana (60) | Cashew Butter (67)<br>Chestnuts (65)<br>Honey Pecans (65)<br>Falafel (62) | Nori Seaweed (67)<br>Bean Chips (62)<br>Sweet Potato Chips (62)<br>Vegetable Chips (60) | Kind Bar (65)<br>Crème Brulée (64)<br>Frozen Yogurt (64)<br>Choc. Pudding (60) | Kashi Cereal (68)<br>Home Granola (67)<br>Grape Nuts (62)<br>Alpen (61) | Whole Grain Pasta (67)<br>Wild Rice (67)<br>Puri Bread (63)<br>Oat Bran Bread (61) | Chocolate Milk (68)<br>Almond Milk Sweet (67)<br>Swiss Cheese (64)<br>No Fat Mozzarella (63) | Chicken Thigh (69)<br>Ground Beef (66)<br>Beef Steak (65)<br>Fried Chicken (62) | Beef & Potatoes (67)<br>Rice w/ vegetables (62)<br>Corn Taco w/ meat (62)<br>Pasta & Sauce (62) | Hummus (69)<br>Mustard (63)<br>Oil (63)<br>Vinegar (60) | Tomato Juice (69)<br>Nonfat Mocha (63)<br>Carrot Juice (60)<br>Boost (60) |
| >=40 | Candied Yams (56)<br>Potato Salad (51)<br>Mashed Potato (46)<br>French Fries (42) | Applesauce (57)<br>Raisins (45)<br>Coconut (42)<br>Cherry Pie Fill. (40) | | Stovetop Popcorn (57)<br>Multigrain Chips (48)<br>Taro Chips (45)<br>Bag Pocorn Plain (43) | Choc. Almonds (59)<br>Choc. Icecream (52)<br>Granola Bar (43)<br>Pumpkin Pie (41) | Bran Flakes (59)<br>Cracklin' Oat Bran (58)<br>Oatmeal Squares (49)<br>Total (48) | Brown Rice (58)<br>French Toast (46)<br>Raisin Bread (44)<br>Multigrain Roll (43) | Rice Milk (58)<br>Mozzarella (47)<br>Goat Cheese (45)<br>Cheddar (41) | Roast Duck (59)<br>Pork Belly (47)<br>Deli Meat (42)<br>Beef Shortribs (40) | Chicken Nachos (56)<br>PB&J Sandwich (54)<br>Flour Taco w/ meat (50)<br>Mac & Cheese (47) | Soy Sauce (57)<br>Miso (51)<br>Fish Sauce (50)<br>Yogurt Dip (43) | Ensure (59)<br>Beer (49)<br>Orange Juice (48)<br>Wine (43) |
| >=20 | Tater Tots (38)<br>Hash Brown (32)<br>Onion Rings (30)<br>Sweet Pickles (25) | Banana chips (38)<br>Dried Mango (38)<br>Dried Cherries (34)<br>Craisens (33) | | Potato Chips (32)<br>Rice Crackers (28)<br>Triscuits (24)<br>Corn Nuts (24) | Sorbet (38)<br>Apple Pie (30)<br>Snickers (24)<br>Cookie (24) | Basic 4 (37)<br>Raisin Bran Crunch (33)<br>Honey Nut Cheerios (27)<br>Wheaties (25) | White bread (37)<br>Pita Bread (27)<br>Pancake (25)<br>Waffle (18) | Brie (39)<br>Feta (36)<br>American (31)<br>Parmesan (27) | Chicken Nuggets (39)<br>Sausage (37)<br>Ham (31)<br>Prosciutto (27) | Cheeseburger (33)<br>Cheese Pizza (36)<br>Grilled Cheese (32)<br>Meet Pizza (26) | Artichoke Dip (34)<br>Mayonnaise (33)<br>Pesto (23)<br>Butter (21) | Red Bull (39)<br>Apple Juice (38)<br>Sweet Tea (35)<br>Grape Juice (34) |
| >=1 | | | | Fritos (19)<br>Pringles (15)<br>Microwave Popcorn (8)<br>Cheetos (1) | Doughnut (19)<br>M&Ms (12)<br>Starbursts (3)<br>Twinkie (1) | Kix (17)<br>Froot Loops (11)<br>Lucky Charms (1)<br>Frosted Flakes (1) | Cheese Bread (15)<br>Scone with Fruit (7)<br>Hush Puppy (3)<br>Biscuit (1) | | Sausage (17)<br>Beef Jerky (2)<br>Bacon (1)<br>Hot Dog (1) | Bologna Sandwich (17)<br>Hot Dog Sandwich (6)<br>Corn Dog (5)<br>Sausage Biscuit (1) | Hollandaise (10)<br>Peanut sauce (10)<br>Alfredo (2)<br>Ketchup (1) | Cola (18)<br>Ginger Ale (15)<br>Daiquiri (6)<br>Vodka (4) |
