## Supplementary figures and images for "A Nutrient Ratio–Based, Web-Enabled Food Quality Score Performs Favorably Compared with Leading Nutrient Profiling Systems for Weight and Blood Pressure"

### Supplemental Figure 2A

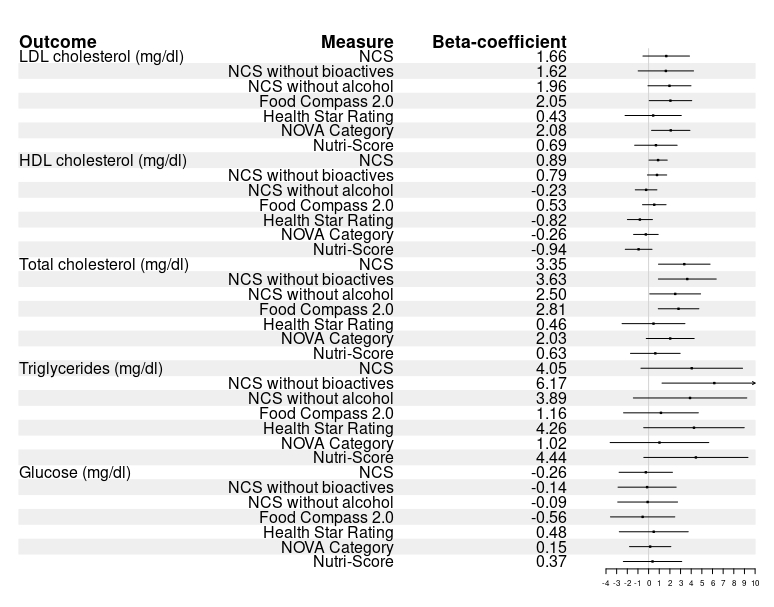

### Supplemental Figure 2B

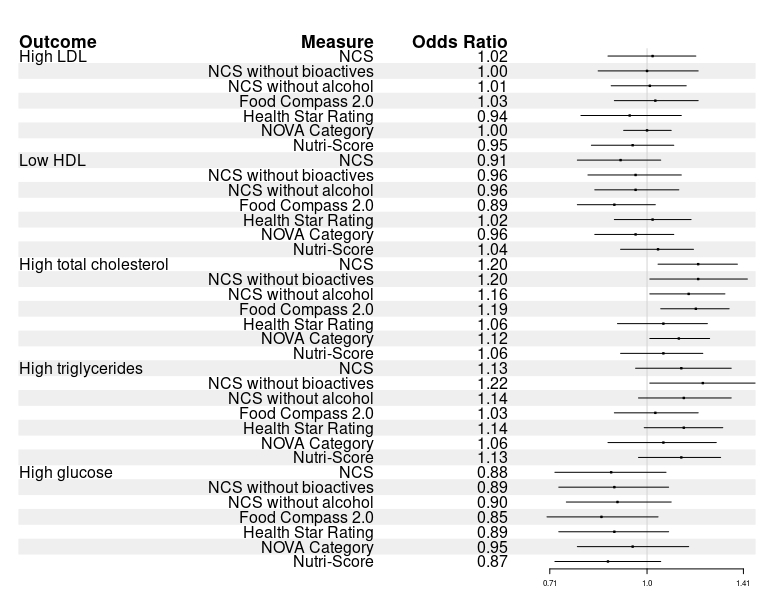

### Supplemental Figure 3A

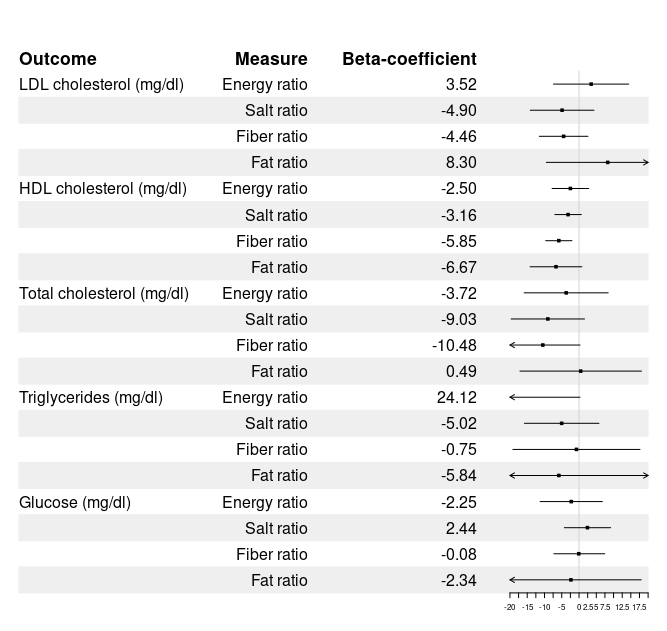

### Supplemental Figure 3B

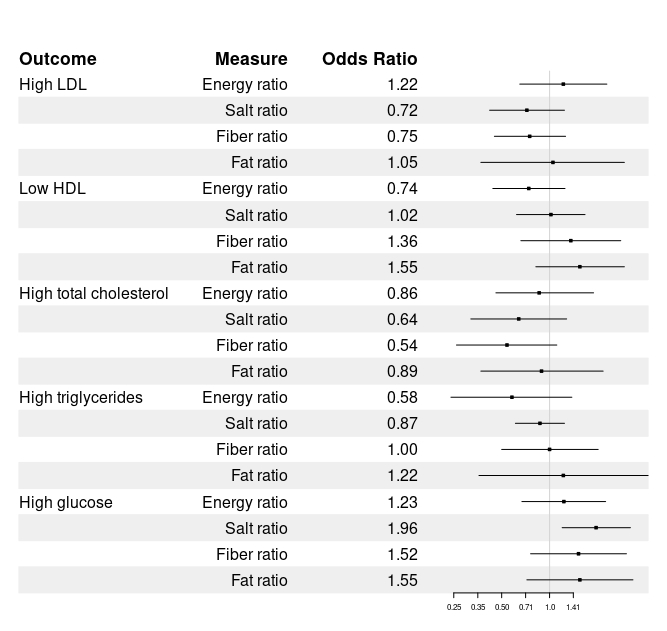
