## Supplemental Figure 4 for "A Nutrient Ratio–Based, Web-Enabled Food Quality Score Performs Favorably Compared with Leading Nutrient Profiling Systems for Weight and Blood Pressure"

| Food category | NCS | NCS, No Bioactives | NCS, No Alcohol | Food Compass 2.0 | Health Star Rating | NOVA Category | Nutri-Score |
|---|---|---|---|---|---|---|---|
| Pizza | -5.49 | -6.11 | -4.57 | -1.47 | -2.50 | -3.02 | -1.97 |
| Soft drinks | -3.51 | -0.99 | -2.32 | -3.56 | -3.37 | -1.05 | -1.09 |
| Frankfurter sandwiches (single code) | -3.29 | -2.40 | -2.95 | -2.51 | -0.88 | -1.68 | -0.90 |
| Egg/breakfast sandwiches (single code) | -2.95 | -1.28 | -2.48 | -0.71 | -0.54 | -0.45 | -1.04 |
| Tea | -1.75 | -0.98 | -1.92 | -2.31 | -2.58 | -0.92 | -1.85 |
| Cookies and brownies | -1.71 | -2.64 | -1.04 | -0.89 | -0.90 | -0.46 | -1.15 |
| Burgers (single code) | -1.11 | -0.37 | -1.09 | -1.87 | -0.60 | -3.81 | -1.28 |
| Liquor and cocktails | -0.98 | -0.44 | -1.84 | -0.95 | -2.61 | -1.71 | -3.48 |
| Biscuits, muffins, quick breads | -0.92 | -1.23 | -0.59 | -0.15 | -0.26 | 0.00 | -0.20 |
| French fries and other fried white potatoes | -0.90 | -0.13 | -0.66 | -0.18 | 0.08 | -1.54 | -0.08 |
| Popcorn | -0.79 | -0.98 | -0.59 | -0.10 | -0.56 | -0.34 | -0.59 |
| Cakes and pies | -0.59 | -0.76 | -0.44 | -0.96 | -0.57 | -0.03 | -0.76 |
| Doughnuts, sweet rolls, pastries | -0.54 | -0.85 | -0.24 | -0.60 | -0.13 | -0.10 | -0.06 |
| Tortilla, corn, other chips | -0.53 | -0.90 | -0.31 | 0.00 | 0.00 | -0.11 | -0.02 |
| Sausages | -0.50 | -0.20 | -0.54 | -0.52 | -0.84 | -0.68 | -1.21 |
| Salad dressings and vegetable oils | -0.49 | -0.59 | -0.61 | -0.02 | -1.19 | -0.41 | -0.71 |
| Chicken/turkey sandwiches (single code) | -0.45 | -0.39 | -0.30 | -0.35 | 0.00 | -1.00 | -0.01 |
| Potato chips | -0.38 | -0.16 | -0.47 | -0.22 | -0.29 | -1.22 | -0.66 |
| Cold cuts and cured meats | -0.32 | -0.25 | -0.48 | -0.38 | -0.53 | -0.56 | -0.37 |
| Rolls and buns | -0.32 | -0.91 | -0.21 | -0.74 | -0.03 | -0.71 | 0.00 |
| Pretzels/snack mix | -0.31 | -0.61 | -0.25 | -0.03 | -0.06 | -0.12 | -0.02 |
| Dark green vegetables, excludes lettuce | -0.30 | -0.11 | -0.40 | -0.88 | -0.15 | -0.27 | -0.25 |
| Chicken patties, nuggets and tenders | -0.27 | -0.21 | -0.19 | -0.13 | -0.04 | -0.46 | -0.34 |
| Candy containing chocolate | -0.23 | -0.23 | -0.11 | -0.16 | -0.58 | -0.38 | -0.24 |
| Crackers, excludes saltines | -0.20 | -0.22 | -0.12 | 0.02 | 0.00 | -0.04 | -0.04 |
| Pancakes, waffles, French toast | -0.19 | -0.20 | -0.10 | -0.03 | 0.03 | -0.02 | 0.02 |
| Burritos and tacos | -0.19 | -0.08 | -0.06 | -0.06 | 0.04 | 0.63 | -0.04 |
| Candy not containing chocolate | -0.19 | -0.21 | -0.29 | -0.13 | -0.17 | -0.28 | -0.24 |
| Sport and energy drinks | -0.16 | -0.23 | -0.26 | -0.40 | -0.88 | -0.04 | -0.23 |
| Spinach | -0.14 | -0.11 | -0.14 | -0.11 | -0.04 | -0.12 | -0.09 |
| Soy-based condiments | -0.14 | -0.03 | -0.16 | -0.11 | -0.21 | -1.06 | -0.22 |
| Mashed potatoes and white potato mixtures | -0.13 | 0.01 | -0.16 | 0.01 | 0.01 | -0.01 | -0.35 |
| Dried fruits | -0.11 | -0.10 | -0.12 | -0.03 | -0.10 | -0.01 | -0.18 |
| Coffee | -0.11 | -0.08 | -0.04 | -0.07 | -0.15 | 0.00 | -0.02 |
| Baby food: fruit | -0.10 | -0.04 | -0.10 | -0.05 | -0.05 | -0.26 | -0.13 |
| Cream cheese, sour cream, whipped cream | -0.09 | -0.10 | -0.13 | -0.11 | -0.33 | -0.10 | -0.17 |
| Not included in a food category | -0.07 | -0.07 | -0.04 | -0.05 | 0.00 | -0.06 | 0.00 |
| Tomatoes | -0.07 | 0.00 | -0.07 | -0.34 | 0.00 | -0.20 | -0.01 |
| Peaches and nectarines | -0.06 | -0.07 | -0.08 | -0.19 | -0.07 | -0.06 | -0.04 |
| Flavored milk, whole | -0.06 | -0.06 | -0.04 | -0.26 | -0.18 | -0.08 | -0.11 |
| Sugars and honey | -0.05 | -0.07 | -0.10 | -0.03 | -0.29 | -0.32 | -0.20 |
| Other Mexican mixed dishes | -0.05 | -0.02 | -0.04 | 0.00 | 0.01 | 1.35 | 0.04 |
| Flavored milk, reduced fat | -0.05 | -0.11 | -0.03 | -0.15 | -0.05 | -0.34 | -0.01 |
| Berries | -0.03 | 0.00 | -0.05 | -0.24 | 0.00 | -0.07 | -0.05 |
| Beef, excludes ground | -0.03 | -0.01 | -0.03 | -0.27 | -0.09 | 0.47 | -0.60 |
| Dips, gravies, other sauces | -0.03 | -0.06 | -0.02 | 0.00 | -0.02 | -0.19 | -0.03 |
| Margarine | -0.02 | -0.01 | -0.02 | -0.01 | -0.02 | 0.01 | 0.00 |
| Turnovers and other grain-based items | -0.02 | -0.03 | -0.01 | 0.01 | 0.02 | 0.26 | -0.01 |
| Shellfish | -0.02 | -0.07 | 0.00 | 0.13 | -0.06 | 0.08 | -0.03 |
| Apple juice | -0.02 | -0.04 | 0.00 | 0.07 | 0.00 | 0.17 | 0.00 |
| Corn | -0.02 | -0.04 | 0.00 | 0.00 | 0.04 | -0.07 | 0.04 |
| Beer | -0.02 | 0.01 | -7.42 | -4.40 | -10.51 | -5.96 | -11.67 |
| Bacon | -0.01 | -0.02 | -0.02 | 0.02 | -0.02 | -0.03 | -0.01 |
| Citrus fruits | -0.01 | -0.04 | -0.02 | -0.05 | -0.08 | 0.00 | -0.10 |
| Fruit drinks | -0.01 | -0.04 | 0.00 | -0.12 | -0.13 | -0.08 | -0.03 |
| Frankfurters | -0.01 | 0.00 | 0.00 | -0.17 | -0.06 | -0.11 | -0.05 |
| Cereal bars | -0.01 | -0.05 | 0.00 | 0.05 | -0.01 | -0.02 | 0.00 |
| Enhanced or fortified water | -0.01 | -0.04 | -0.01 | -0.05 | -0.03 | -0.01 | 0.00 |
| Cheese sandwiches (single code) | -0.01 | -0.14 | 0.00 | 0.00 | -0.20 | -0.05 | -0.14 |
| Other vegetables and combinations | 0.00 | 0.01 | 0.00 | 0.00 | 0.20 | -0.02 | 0.16 |
| Carrots | 0.00 | 0.01 | -0.01 | -0.07 | 0.04 | -0.05 | 0.02 |
| Nachos | 0.00 | 0.00 | -0.01 | -0.04 | -0.05 | -0.08 | -0.02 |
| Milk, nonfat | 0.00 | 0.00 | 0.00 | -0.01 | 0.01 | 0.00 | 0.03 |
| Vegetable dishes | 0.00 | -0.03 | 0.00 | 0.03 | -0.01 | 0.01 | -0.01 |
| Mustard and other condiments | 0.00 | 0.00 | 0.00 | -0.10 | 0.05 | -0.08 | 0.02 |
| Broccoli | 0.00 | -0.01 | 0.02 | 0.06 | 0.09 | 0.01 | 0.17 |
| Bagels and English muffins | 0.00 | -0.22 | 0.00 | -0.17 | 0.04 | -0.32 | 0.10 |
| Jams, syrups, toppings | 0.00 | 0.00 | 0.00 | 0.02 | 0.00 | 0.00 | -0.02 |
| Grits and other cooked cereals | 0.00 | -0.01 | 0.00 | -0.06 | 0.00 | 0.00 | -0.02 |
| Cabbage | 0.00 | 0.00 | 0.00 | -0.05 | 0.00 | 0.00 | 0.00 |
| Seafood sandwiches (single code) | 0.00 | 0.00 | 0.00 | 0.00 | 0.00 | 0.00 | 0.00 |

| | | | | | | | |
|---|---|---|---|---|---|---|---|
| Blueberries and other berries | 0.00 | 0.00 | 0.00 | 0.00 | 0.00 | 0.00 | 0.00 |
| Pears | 0.00 | 0.00 | 0.00 | 0.00 | 0.00 | 0.00 | 0.00 |
| Pineapple | 0.00 | 0.00 | 0.00 | 0.00 | 0.00 | 0.00 | 0.00 |
| Vegetables on a sandwich | 0.00 | 0.00 | 0.00 | 0.00 | 0.00 | 0.00 | 0.00 |
| Tap water | 0.00 | 0.00 | 0.00 | 0.00 | 0.00 | 0.00 | 0.00 |
| Bottled water | 0.00 | 0.00 | 0.00 | 0.00 | 0.00 | 0.00 | 0.00 |
| Baby food: vegetable | 0.00 | 0.00 | 0.00 | 0.00 | 0.00 | 0.00 | 0.00 |
| Baby food: meat and dinners | 0.00 | 0.00 | 0.00 | 0.00 | 0.00 | 0.00 | 0.00 |
| Baby food: yogurt | 0.00 | 0.00 | 0.00 | 0.00 | 0.00 | 0.00 | 0.00 |
| Baby food: snacks and sweets | 0.00 | 0.00 | 0.00 | 0.00 | 0.00 | 0.00 | 0.00 |
| Baby juice | 0.00 | 0.00 | 0.00 | 0.00 | 0.00 | 0.00 | 0.00 |
| Baby water | 0.00 | 0.00 | 0.00 | 0.00 | 0.00 | 0.00 | 0.00 |
| Formula, ready-to-feed | 0.00 | 0.00 | 0.00 | 0.00 | 0.00 | 0.00 | 0.00 |
| Formula, prepared from powder | 0.00 | 0.00 | 0.00 | 0.00 | 0.00 | 0.00 | 0.00 |
| Formula, prepared from concentrate | 0.00 | 0.00 | 0.00 | 0.00 | 0.00 | 0.00 | 0.00 |
| Human milk | 0.00 | 0.00 | 0.00 | 0.00 | 0.00 | 0.00 | 0.00 |
| Saltine crackers | 0.00 | -0.01 | 0.00 | 0.00 | 0.01 | 0.00 | 0.04 |
| Gelatins, ices, sorbets | 0.00 | 0.00 | 0.03 | 0.01 | 0.00 | 0.00 | 0.00 |
| Sugar substitutes | 0.00 | 0.00 | -0.01 | -0.02 | -0.09 | 0.25 | -0.03 |
| Nutrition bars | 0.00 | 0.00 | 0.00 | 0.04 | -0.04 | -0.03 | -0.03 |
| Cheese | 0.00 | -0.10 | 0.01 | 0.07 | -0.36 | -0.02 | -0.08 |
| Liver and organ meats | 0.00 | 0.02 | 0.00 | 0.07 | 0.00 | 0.08 | 0.00 |
| Diet soft drinks | 0.00 | 0.00 | 0.00 | -0.02 | -0.13 | 0.28 | 0.00 |
| Other diet drinks | 0.00 | 0.03 | 0.00 | 0.00 | -0.01 | -0.07 | -0.01 |
| Wine | 0.00 | 0.00 | -0.60 | -0.07 | -0.76 | -0.30 | -1.13 |
| Fried vegetables | 0.00 | 0.00 | 0.01 | 0.01 | 0.00 | -0.01 | 0.00 |
| Fried rice and lo/chow mein | 0.01 | 0.00 | 0.01 | 0.02 | 0.42 | 3.04 | 0.52 |
| String beans | 0.01 | 0.01 | 0.00 | -0.05 | -0.01 | -0.04 | 0.00 |
| Protein and nutritional powders | 0.01 | 0.00 | 0.01 | 0.06 | -0.04 | -0.01 | -0.06 |
| Other red and orange vegetables | 0.01 | 0.00 | 0.00 | 0.00 | 0.00 | 0.01 | 0.00 |
| Other sandwiches (single code) | 0.01 | 0.00 | 0.02 | -0.16 | 0.02 | -0.57 | 0.07 |
| Other dark green vegetables | 0.01 | 0.03 | 0.00 | 0.00 | 0.00 | 0.10 | 0.03 |
| Milk substitutes | 0.01 | 0.03 | 0.00 | -0.04 | -0.01 | 0.00 | 0.01 |
| Other fruit juice | 0.01 | 0.02 | 0.01 | 0.39 | 0.01 | 0.45 | 0.00 |
| Bean, pea, legume dishes | 0.02 | 0.01 | 0.00 | 0.00 | 0.03 | -0.01 | 0.03 |
| Egg rolls, dumplings, sushi | 0.02 | -0.03 | 0.01 | 0.00 | -0.01 | 0.25 | 0.00 |
| Ready-to-eat cereal, higher sugar (>21.2g/100g) | 0.02 | 0.00 | 0.02 | 0.44 | 0.09 | 0.02 | 0.04 |
| Diet sport and energy drinks | 0.02 | 0.02 | 0.01 | 0.00 | 0.01 | 0.00 | 0.02 |
| Vegetable juice | 0.02 | 0.08 | 0.03 | 0.11 | 0.00 | 0.00 | 0.00 |
| Other fruits and fruit salads | 0.02 | 0.02 | 0.00 | 0.01 | 0.02 | 0.00 | 0.03 |
| Butter and animal fats | 0.03 | 0.01 | 0.09 | 0.11 | 0.05 | 0.07 | 0.02 |
| White potatoes, baked or boiled | 0.03 | 0.00 | 0.03 | 0.10 | 0.03 | 0.33 | 0.00 |
| Flavored milk, nonfat | 0.04 | 0.04 | 0.13 | 0.03 | 0.22 | 0.01 | 0.37 |
| Processed soy products | 0.05 | 0.01 | 0.04 | 0.07 | 0.02 | -0.07 | 0.05 |
| Peanut butter and jelly sandwiches (single code) | 0.05 | 0.00 | 0.10 | 0.05 | 0.01 | -0.19 | 0.00 |
| Flavored or carbonated water | 0.05 | 0.09 | 0.02 | -0.02 | 0.01 | -0.02 | 0.02 |
| Citrus juice | 0.05 | 0.05 | 0.06 | 0.74 | 0.18 | 1.09 | 0.01 |
| Turkey, duck, other poultry | 0.05 | 0.10 | 0.06 | 0.19 | 0.27 | 0.11 | 0.07 |
| Baby food: cereals | 0.06 | 0.08 | 0.06 | 0.02 | 0.12 | -0.01 | 0.14 |
| Melons | 0.06 | 0.03 | 0.02 | 0.29 | 0.00 | 0.08 | 0.01 |
| Coleslaw, non-lettuce salads | 0.06 | 0.04 | 0.02 | 0.04 | 0.03 | 0.05 | 0.03 |
| Flavored milk, lowfat | 0.06 | 0.07 | 0.07 | -0.06 | 0.05 | -0.17 | 0.05 |
| Lamb, goat, game | 0.06 | 0.04 | 0.09 | 0.14 | 0.04 | 0.32 | 0.05 |
| Macaroni and cheese | 0.07 | 0.14 | 0.07 | 0.01 | 0.03 | 0.02 | -0.01 |
| Milk shakes and other dairy drinks | 0.07 | 0.17 | 0.13 | -0.03 | -0.02 | -0.14 | 0.14 |
| Mango and papaya | 0.07 | 0.04 | 0.06 | 0.15 | 0.06 | 0.08 | 0.11 |
| Nutritional beverages | 0.09 | 0.12 | 0.08 | 0.11 | -0.13 | 0.00 | -0.02 |
| Pasta, noodles, cooked grains | 0.09 | 0.02 | 0.03 | 0.20 | 0.03 | 0.70 | 0.15 |
| Pork | 0.09 | 0.12 | 0.04 | 0.01 | -0.05 | 0.21 | -0.23 |
| Pasta mixed dishes, excludes macaroni and cheese | 0.09 | 0.15 | 0.12 | 0.23 | 0.38 | 0.45 | 1.45 |
| Cream and cream substitutes | 0.10 | 0.06 | 0.08 | 0.01 | 0.00 | 0.01 | 0.13 |
| Grapes | 0.12 | 0.06 | 0.08 | 0.10 | 0.06 | 0.01 | 0.13 |
| Ice cream and frozen dairy desserts | 0.13 | 0.05 | 0.29 | -0.36 | -0.03 | -0.35 | -0.07 |
| Lettuce and lettuce salads | 0.14 | 0.49 | 0.20 | -0.01 | 0.97 | 0.11 | 0.45 |
| Ground beef | 0.15 | 0.14 | 0.13 | 0.04 | -0.03 | 0.19 | -0.07 |
| Milk, lowfat | 0.17 | 0.16 | 0.16 | 0.10 | 0.22 | 0.02 | 0.33 |
| Apples | 0.17 | 0.17 | 0.11 | 0.13 | 0.05 | 0.00 | 0.02 |
| Onions | 0.17 | 0.23 | 0.14 | -0.01 | 0.07 | 0.07 | 0.09 |
| Chicken, whole pieces | 0.18 | 0.32 | 0.06 | 0.19 | 0.00 | 0.05 | -0.23 |
| Pasta sauces, tomato-based | 0.20 | 0.34 | 0.14 | 0.23 | 0.13 | 0.06 | 0.01 |
| Mayonnaise | 0.20 | 0.24 | 0.16 | 0.32 | 0.05 | 0.32 | 0.03 |
| Poultry mixed dishes | 0.24 | 0.38 | 0.31 | 0.17 | 0.28 | 0.04 | 0.41 |
| Ready-to-eat cereal, lower sugar (=<21.2g/100g) | 0.24 | 0.28 | 0.30 | 0.58 | 0.57 | -0.09 | 0.57 |

| | | | | | | | |
|---|---|---|---|---|---|---|---|
| Vegetable mixed dishes | 0.24 | 0.26 | 0.22 | 0.36 | 0.19 | 0.16 | 0.04 |
| Cottage/ricotta cheese | 0.25 | 0.20 | 0.22 | 0.10 | 0.13 | 0.00 | 0.06 |
| Meat mixed dishes | 0.26 | 0.33 | 0.33 | 0.01 | 0.83 | 0.42 | 0.63 |
| Milk, reduced fat | 0.28 | 0.20 | 0.54 | 0.51 | 0.68 | 0.98 | 0.66 |
| Yogurt, regular | 0.30 | 0.23 | 0.28 | 0.03 | 0.41 | 0.00 | 0.09 |
| Yogurt, whole and reduced fat | 0.35 | 0.36 | 0.34 | 0.49 | 0.19 | 0.19 | 0.16 |
| Pudding | 0.36 | 0.25 | 0.38 | 0.11 | 0.17 | 0.31 | 0.09 |
| Tortillas | 0.41 | 0.61 | 0.44 | 0.19 | 1.21 | 0.08 | 1.32 |
| Yogurt, lowfat and nonfat | 0.45 | 0.30 | 0.49 | 0.14 | 0.47 | -0.05 | 0.38 |
| Other starchy vegetables | 0.46 | 0.08 | 0.39 | 0.64 | 0.46 | 0.54 | 0.20 |
| Milk, whole | 0.49 | 0.30 | 0.60 | 0.76 | 0.21 | 0.93 | 0.44 |
| Tomato-based condiments | 0.50 | 0.20 | 0.83 | 0.53 | 0.68 | 0.33 | 1.29 |
| Olives, pickles, pickled vegetables | 0.59 | 0.67 | 0.52 | 0.51 | 0.56 | 0.34 | 0.61 |
| Eggs and omelets | 0.70 | 0.69 | 0.88 | 1.92 | 0.42 | 0.36 | 0.06 |
| Smoothies and grain drinks | 0.72 | 0.76 | 0.92 | 2.33 | -0.05 | 1.64 | 0.00 |
| Yogurt, Greek | 0.92 | 0.73 | 0.87 | 0.77 | 0.72 | 0.00 | 0.61 |
| Soups | 1.05 | 0.88 | 1.27 | 0.81 | 2.05 | 1.15 | 2.03 |
| Stir-fry and soy-based sauce mixtures | 1.09 | 0.55 | 0.86 | 0.58 | 0.18 | 1.57 | 0.33 |
| Seafood mixed dishes | 1.09 | 0.71 | 1.07 | 1.31 | 0.31 | 0.12 | 0.45 |
| Yeast breads | 1.10 | 0.15 | 1.21 | 1.09 | 1.79 | 0.02 | 2.65 |
| Bananas | 1.13 | 0.64 | 1.09 | 1.42 | 0.84 | 1.13 | 1.03 |
| Rice mixed dishes | 1.96 | 0.90 | 2.10 | 1.24 | 1.71 | 1.25 | 1.60 |
| Fish | 2.52 | 1.23 | 2.34 | 4.60 | 0.68 | 1.63 | 0.66 |
| Oatmeal | 3.22 | 2.20 | 3.26 | 3.33 | 1.70 | 2.04 | 3.01 |
| Nuts and seeds | 4.53 | 3.40 | 4.27 | 9.81 | 3.70 | 1.67 | 1.14 |
| Beans, peas, legumes | 5.91 | 5.26 | 5.98 | 4.36 | 5.44 | 1.64 | 5.66 |
| Rice | 6.14 | 2.42 | 6.51 | 4.70 | 5.39 | 12.89 | 6.90 |
